# Adaptation of the multiplexed CRISPR-Cas13 CARMEN RVP assay for longitudinal detection of respiratory pathogens from air samples

**DOI:** 10.1101/2025.10.21.25338488

**Authors:** Amy L. Ellis, Miranda Stauss, Patrick Barros Tiburcio, Isla E. Emmen, Paul T. Edlefsen, Ewelina Kosmider, Shari Barlow, Maureen Goss, Jonathan L. Temte, Elyse Stachler, Kyle McMahon, Pardis Sabeti, David H. O’Connor, Shelby L. O’Connor

## Abstract

Air sampling is a non-invasive alternative to individual testing for respiratory pathogens. Alternative methods to the “gold standard” quantitative RT-PCR (qRT-PCR) are required to enable higher throughput, lower cost, and more multiplexed detection of pathogens. The multiplexed CRISPR-Cas13 CARMEN Respiratory Viral Panel (RVP) was described previously for high-throughput detection of nine respiratory pathogens from nasal swab samples. Here, we modified and optimized the CARMEN RVP assay to overcome the unique challenges of air samples, including low biomass and environmental inhibitors. We monitored for SARS-CoV-2 and influenza A (Flu A) via qRT-PCR in air samples from 15 schools within Dane County, Wisconsin (USA) during the 2023-2024 school year. SARS-CoV-2 was detectable throughout the entire sampling period, while Flu A detection was seasonal from November 2023 to March 2024. We then analyzed a subset of samples from seven schools using an optimized CARMEN RVP assay for air surveillance (RVP_air) and compared results to qRT-PCR. The RVP_air assay detected several additional pathogens beyond our primary targets. The frequencies and patterns of SARS-CoV-2 positivity, but not Flu A, were similar between qRT-PCR and RVP_air across the 2023-2024 sampling period. We developed a secondary panel (RVP_air_flu) to better detect both H1N1 and H3N2 subtypes. Finally, we compared air sample results to clinical nasal swabs collected from the same school district. For several pathogens (SARS-CoV-2, HCoV-OC43, Flu A), positive air detections coincided with positive nasal swabs. These findings demonstrate that the RVP_air assay can effectively detect airborne pathogens from infected individuals within indoor spaces.

**IMPORTANCE:** Air sampling offers a cost-effective alternative to individual testing for respiratory pathogens within congregate settings. Optimization and use of multi-pathogen assays are especially valuable for capturing the breadth of pathogens that may be present simultaneously in the same space. The modified CARMEN RVP assays (RVP_air and RVP_air_flu) detected SARS-CoV-2 and Flu A during similar sampling time periods compared to qRT-PCR, while also detecting several additional respiratory pathogens (seasonal Coronaviruses, Respiratory Syncytial Virus). Importantly, pathogens detected from air samples corresponded to those detected from nasal swabs collected from individuals in the same spaces. Together, these findings highlight the utility of the RVP_air and RVP_air_flu assays as alternatives to qRT-PCR for environmental surveillance, with applications extending to other congregate spaces (hospitals, long-term care facilities) and high-risk settings, better informing communities and improving public health.

## INTRODUCTION

Virus surveillance via nasal swab testing in schools provides an indication of ongoing virus transmission in the surrounding communities (1). However, clinical testing for SARS-CoV-2 in schools is cumbersome and expensive (2–4). We and others have found that viral RNA from respiratory pathogens can be routinely detected in indoor air samples and correlate with clinically confirmed cases in the same space (5–7).

Pathogen detection from air samples can be challenging. Air samples can contain dust, dirt, particulates, and inhibitors that can interfere with enzymatic reactions during PCR (8). The biomass of air samples is also low (9), so highly sensitive and specific assays are needed to detect the low concentration of nucleic acids present in a small volume of the eluted air samples. Therefore, assays detecting genetic material collected from air samples must be robust enough to be able to overcome these challenges.

Air samples can contain nucleic acids from many different viruses, particularly during peaks in respiratory virus seasons. Therefore, assessing and improving the performance of assays that detect many pathogens from a single sample is necessary. Assays such as qRT-PCR and digital PCR (dPCR) are highly sensitive and can be multiplexed, but most platforms can only detect 1-4 targets per sample. Multi-pathogen qRT-PCR detection from air samples has been achieved (5, 10–11), but the assays used in these studies utilized multiple primer pools or expensive assay systems, making their use prohibitive either monetarily or in terms of use of sample volume.

We sought to improve the detection of multiple pathogens in a more cost-effective manner from collected air samples using a CRISPR/Cas13-based PCR technology that leverages highly specific RNA-targeting of the Cas13 enzyme in conjunction with nucleic acid amplification techniques (12–13). The CRISPR/Cas13 system enables precise and sensitive detection of target RNA with minimal cross-reactivity (12, 14). The Combinatorial Arrayed Reactions for Multiplexed Evaluation of Nucleic acids Respiratory Virus Panel (CARMEN RVP) assay was developed during the COVID-19 pandemic and allowed for high-throughput, sensitive detection of multiple respiratory pathogens from clinical swabs (15). The assay is run on the Biomark X platform and has the potential to assess for the presence of up to 24 different targets from 192 different samples. We calculated the cost to be approximately $10-20/sample, and only 10μL of sample is used per test. The combination of minimal use of sample volume, low cost, and high throughput makes the CARMEN RVP assay an attractive alternative to other multiplex platforms. However, this assay has not previously been tested for its performance with environmental samples.

## Results

### The development of a modified CARMEN panel (RVP_air) facilitates pathogen detection from air samples

#### Initial testing of original CARMEN RVP with air samples

The CARMEN (RVP) assay was originally designed for high-throughput detection of nine respiratory pathogens in clinical patient samples (15). We initially tested how the CARMEN RVP assay would perform with environmental samples using RNA extracted from 107 air samples from schools previously tested via qRT-PCR for SARS-CoV-2 and Flu A genetic material, as well as RNAseP, which is used as a positive control for the presence of human genetic material that indicates successful air collection. Of these 107 samples, 98 were also positive for human control RNAseP via CARMEN RVP. Of those 98 positive samples, we excluded any inconclusive samples (samples where only one of two replicates was positive) for qRT-PCR. We then performed kappa concordance tests to determine agreement between qRT-PCR and CARMEN RVP results (Fig. 1A). Kappa scores near zero are considered poor, 0-0.2 slight, 0.2-0.4 fair, 0.4-0.6 moderate, 0.6-0.8 substantial, and 0.8-1 excellent (16). We observed very poor concordance between qRT-PCR and CARMEN RVP when examining SARS-CoV-2 (1.3% positive sample agreement, 8.6% agreement all samples; k=0.002) and Flu A (k=0.000) (Fig. 1A). The Flu A data appeared to have high agreement when considering all samples (91.7%), but this was skewed heavily due to the majority of samples being negative for Flu A via qRT-PCR. There were no Flu A positive samples via CARMEN RVP and therefore no positive agreements (Fig. 1A). Finally, there were no other pathogens detected by the CARMEN RVP assay in any of the 98 samples (data not shown). These initial data suggested that improvements to the CARMEN RVP assay were needed for air samples.

**Fig. 1.**
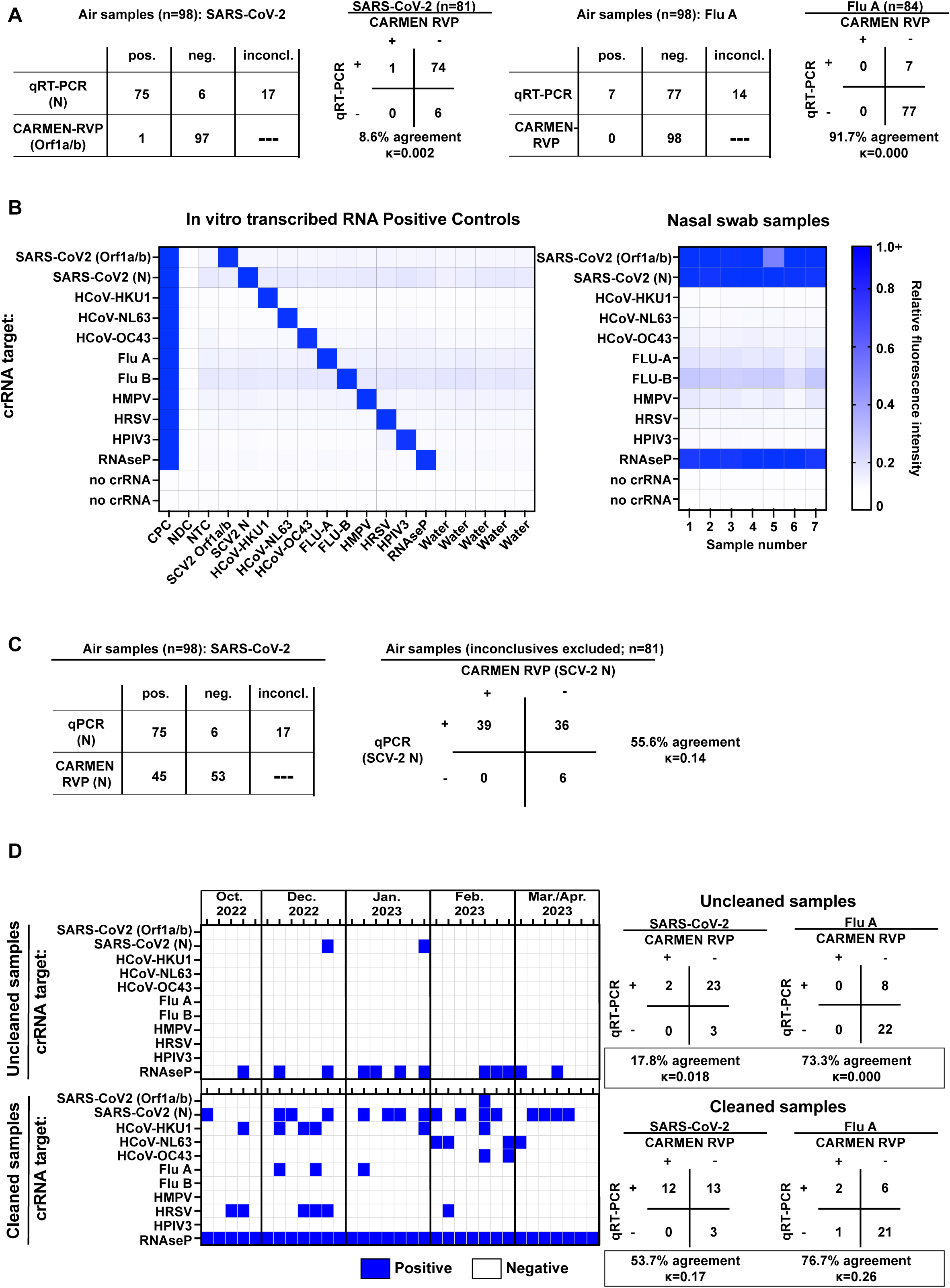
Optimization experiments to adapt CARMEN RVP for use with air samples. A, The CARMEN RVP assay as initially described in (15) was performed on RNA isolated from 98 air samples, and results for SARS-CoV-2 (left) and Flu A (right) were compared to results obtained from qRT-PCR for the same samples. Total positive, negative, and inconclusive results are shown in the tables on the left for the indicated targets for each assay, and one-to-one comparisons and kappa score statistics are shown in the tables on the right. B, The CARMEN RVP assay was modified to include a target for SARS-CoV-2 N, and the new assay, termed RVP_air, was tested on positive control RNA (left) and nasal swabs (right) to confirm the new SARS-CoV-2 N target did not cross-react with the original CARMEN RVP targets. Heatmap colors indicate relative FAM fluorescence intensity of the fluorescent probe used for assay readout. C, RVP_air assay was performed on the same RNA from air samples indicated in A. Positive hits for SARS-CoV-2 N for both RVP_air and qRT-PCR were compared. The table on the left indicates total positive, negative, and inconclusive results, and the table on the right shows one-to-one comparisons performed for kappa concordance tests. D, RNA was isolated from air samples collected from the indicated timepoints from October 2022 to April 2023. The RNA samples were divided in half, and half of the volume was cleaned using a OneStep PCR inhibitor removal kit as indicated in the methods. RVP_air was performed on uncleaned (top) and cleaned (bottom) samples. Positive hits are in blue, and negative results are in white. The tables on the right show kappa concordance test comparisons between uncleaned (top heat map) and uncleaned (bottom heat map) RVP_air results and qRT-PCR results that were previously described in(6) for SARS-CoV-2 and Flu A.

#### Modification of the CARMEN RVP assay to detect the SARS-CoV-2 N gene

The primers used in our qRT-PCR assay to amplify SARS-CoV-2 targeted the nucleocapsid (N) gene, which is one of the most abundant transcripts produced during viral infection (17–19), whereas the CARMEN RVP assay was designed with a crRNA targeting Orf1a/b, which is typically present at a lower abundance during viral infections than the N gene (18–19). We used the Activity-informed Design with All-inclusive Patrolling of Targets (ADAPT) program (20) to design a crRNA and primer set for the SARS-CoV-2 N gene. Using positive controls for each target, the SARS-CoV-2 N-specific primers and crRNA did not cross-react with any of the targets in the original CARMEN RVP assay (Fig. 1B). Using seven SARS-CoV-2+ clinical swab samples, we successfully detected RNAseP and SARS-CoV-2 targets with the modified CARMEN RVP.

We then applied the modified CARMEN RVP to the 98 air samples examined in Fig. 1A. While the SARS-CoV-2 Orf1a/b target was only observed in one sample (Fig. 1A), SARS-CoV-2 N was detected in 45 total samples (Fig. 1C). We repeated the kappa concordance test between the qRT-PCR and CARMEN RVP assays on the subset of 81 samples after excluding the 17 inconclusive samples. Of those remaining 81 samples, 39 were positive for SARS-CoV-2 via CARMEN RVP (52% sample agreement among positive samples, 55.6% sample agreement across all samples). While adding the nucleocapsid target increased the kappa concordance score to 0.14, no other pathogens were detected, potentially indicating that additional improvements were still needed for the CARMEN RVP assay.

#### Removal of PCR inhibitors from air sample RNA improves multi-pathogen detection from air samples

The original CARMEN RVP was designed for use with clinical nasal swabs (15), but environmental samples, such as those collected from air and water, contain contaminants and PCR inhibitors (10). PCR inhibitor removal kits are frequently used in processing samples collected from water (21, 22), so we applied a similar strategy to air samples. We isolated RNA from 33 additional air samples previously tested via qRT-PCR for Flu A and SARS-CoV-2 (6). One-half of each RNA isolate sample was left uncleaned, while the other half was applied to a PCR inhibitor removal column to remove environmental contaminants (Zymo Research; see methods). CARMEN RVP was performed on uncleaned and cleaned samples. In contrast to the uncleaned samples, all 33 cleaned samples were positive for RNAseP, 16 samples were positive for SARS-CoV-2, and additional pathogens were now detected (HCoV-HKU1, -NL63, and -OC43; Flu A; and HRSV; Fig. 1D). When cleaned and uncleaned datasets were compared to the samples that were positive for SARS-CoV-2 and Flu A via qRT-PCR (qRT-PCR results originally described in (6)), CARMEN RVP exhibited improved performance after environmental inhibitor removal. When <u>only</u> considering samples that were positive for SARS-CoV-2 (n=25) and Flu A (n=8) via qRT-PCR, the percent detection of SARS-CoV-2 relative to qRT-PCR increased from 8% (uncleaned) to 48% (cleaned), and detection of Flu A increased from 0% (uncleaned) to 25% (cleaned). When considering agreement between all samples (positive and negative), the concordance between CARMEN RVP and qRT-PCR showed improvement for both SARS-CoV-2 and Flu A (17.8% to 53.7% agreement, κ=0.018 to 0.17 for SARS-CoV-2; 73.3% to 76.7% agreement, κ=0.000 to 0.26 for Flu A; Fig. 1D). The detection of additional targets indicated that removal of dirt and contaminants did improve overall assay performance. The detection of SARS-CoV-2, Flu A, and HRSV in the CARMEN RVP corresponded to historical peaks in emergency room visits related to these pathogens in Dane County, Wisconsin (https://publichealthmdc.com/your-health/respiratory-illness/dashboard). We term this improved assay “RVP_air”.

#### Detection of SARS-CoV-2 and Flu A nucleic acids in the air of K-12 schools in 2023-2024 by qRT-PCR

We collected air samples from high-traffic areas of 15 K-12 schools in Dane County, Wisconsin, from August 2023 to June 2024. During the course of the academic year, collected air was tested for the presence of SARS-CoV-2 and Flu A genetic material using qRT-PCR assays for all 15 schools (Fig. 2). SARS-CoV-2 genetic material was detected continuously, with at least one positive sample per week, for the majority of the 2023/2024 school year. The highest percentage of positive samples for SARS-CoV-2 was observed from December 2023 through February 2024, which corresponded to the most significant peak in detection of SARS-CoV-2 from wastewater in the Madison, WI area (https://www.dhs.wisconsin.gov/covid-19/wastewater.htm; peak near 12/31/23). Flu A genetic material was only detected from November 2023 to April 2024. The weeks with peak frequencies of Flu A-positive samples were observed from December 2023 to January 2024. The presence of Flu A in the schools corresponded to when the peak Flu A activity was reported for the Dane County area (https://publichealthmdc.com/your-health/respiratory-illness/dashboard).

**Fig. 2.**
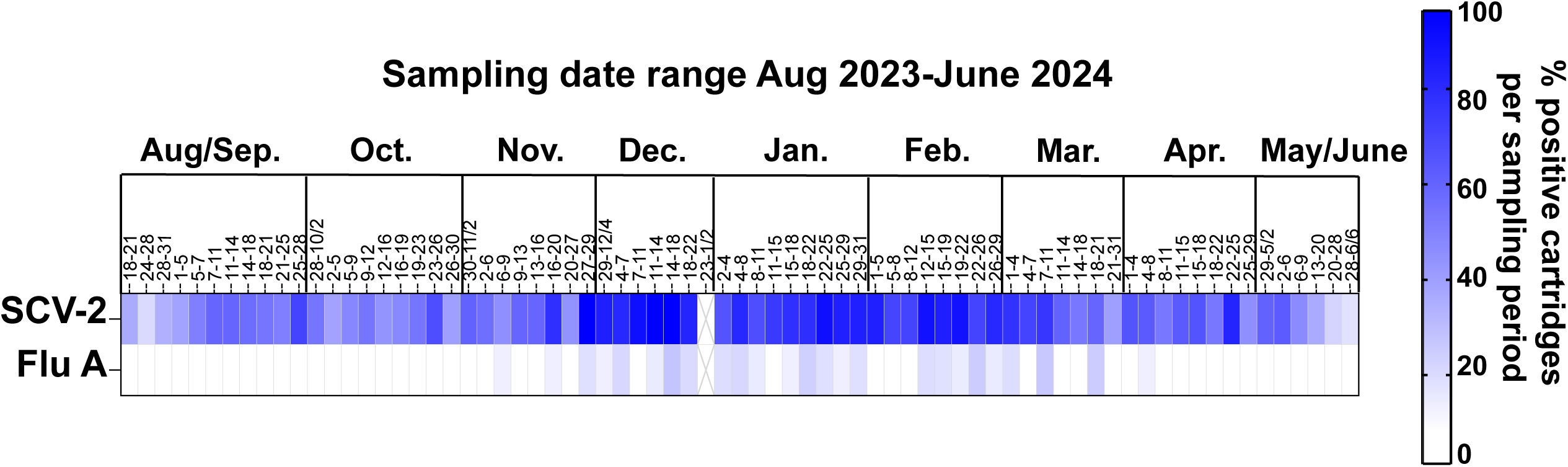
Quantitative reverse transcription PCR (qRT-PCR) results for SARS-CoV-2 and Flu A from 15 K-12 schools in Dane County, WI, for the 2023-2024 school year. Quantitative RT-PCR (qRT-PCR) was performed on RNA isolated from air samples collected from August 2023 to June 2024 from 15 K-12 schools. SARS-CoV-2 and Flu A positivity were measured as indicated in the methods, and the percentage of positive air cartridges per week is shown in the heatmap. Dark blue color indicates a higher frequency of positive air cartridges.

### The RVP_air panel detects seasonal respiratory pathogens present in the air of K-12 schools in 2023-2024, previously not tested by qRT-PCR

We applied the RVP_air assay to a subset of samples from seven of the K-12 schools tested in 2023/2024 (Fig. 3A). We included 347 samples such that there were 5 to 7 samples characterized throughout most weeks of the 2023-2024 school year. We generated a heat map of the frequency of schools positive for a specific pathogen in a given week (Fig. 3A). Using the RVP_air assay, we detected genetic material from additional pathogens throughout the school year that we previously did not measure via qRT-PCR. For example, HCoV-HKU1 and HCoV-OC43 coronaviruses were observed from January through March, and HCoV-NL63 coronavirus was detected from March through May (Fig. 3A). HRSV was most frequently detected in air samples collected in January and February (Fig. 3A).

**Fig. 3.**
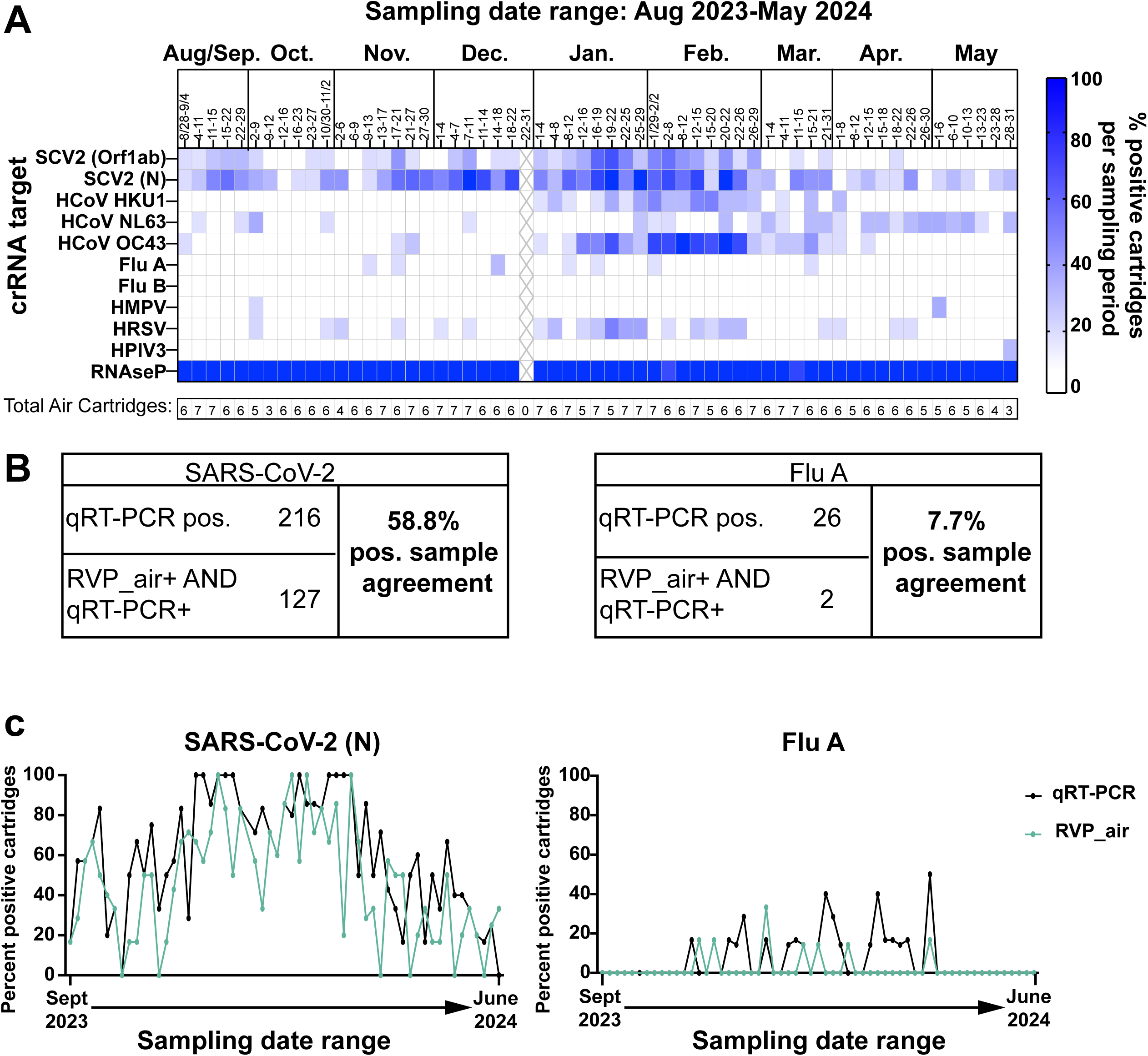
Comparative analyses between RVP_air and qRT-PCR on RNA collected from seven K-12 schools from the 2023/2024 school year. A, RVP_air assay was performed on air samples collected from August 2023 to June 2024 from seven of the 15 K-12 schools shown in Fig. 2. The heatmap indicates the percent of positive samples for each indicated pathogen listed on the left for the sampling date range. The “X” indicates winter break. B, The tables show the total positive samples for SARS-CoV-2 (left) and Flu A (right) for qRT-PCR and RVP_air. The table also shows the number of samples that were positive for both assays for the indicated pathogens (bottom rows) which were used to calculate the percentage of positive sample agreement. C, Comparative analyses were performed between the percent of samples positive for either SARS-CoV-2 (left graph) or Flu A (right graph) for either RVP_air (green lines) and qRT-PCR (black lines) for air samples collected longitudinally from the subset of seven schools shown in A.

We calculated the percentage of samples that were positive for SARS-CoV-2 via RVP_air relative to the number of samples that were positive for SARS-CoV-2 via qRT-PCR. Of 216 samples positive for SARS-CoV-2 via qRT-PCR, 58.8% (127) were also positive via RVP_air (Fig. 3B, Supplementary Table 1). We also performed kappa concordance tests to compare the one-to-one agreement of all samples (both positive and negative) for SARS-CoV-2 and Flu A by qRT-PCR and RVP_air (Supplementary Table 2). For SARS-CoV-2, the kappa score ranged from 0.104 to 0.372. To assess whether these scores exceeded what would be expected by chance alone, we performed a permutation test to obtain p-values. All results were significant, indicating that even the lowest kappa score was unlikely to have occurred by chance. The agreement between qRT-PCR and RVP_air was greatest for SARS-CoV-2 when either Orf1ab or N gene targets were positive for RVP_air, and at least one replicate for qRT-PCR had a Ct value of 35 or lower (kappa=0.372, p-value<0.001). It is logical that the concordance was higher for qRT-PCR results associated with lower Ct values, as it indicates that more genetic material was likely present and therefore increases the likelihood for detection by multiple different assays.

For Flu A sample-to-sample concordance, we observed that of the samples that were positive via qRT-PCR (26 total samples), only 2 of those 26 were also positive for Flu via RVP_air (Fig. 3B, Supplementary Table 1). Furthermore, when comparing kappa concordance for all samples (inclusive of both positive and negative results), no concordance was observed between RVP_air and qRT-PCR, with kappa scores ranging from -0.029 to 0.085, with no statistically significant observations. We partially attributed the lack of statistical significance to the very low numbers of samples that tested positive for Flu A using qRT-PCR and RVP_air for this sample set. This observation raised concerns about assay performance for Flu A.

Next, we evaluated whether the percentage of positive cartridges detected by qRT-PCR and RVP_air followed the same trends over time using Spearman correlation (Fig. 3C, Supplementary Table 2). For SARS-CoV-2, the results were consistent with the kappa score analysis, with all correlations being significant. The highest correlations were observed when considering either gene N alone (Spearman r=0.705, p-value <0.001) or gene N and Orf1ab combined (Spearman r=0.728, p-value <0.001), using a positivity threshold of at least one replicate with a Ct value of 35 or lower. For Flu A, the correlation between the percent of positive cartridges between RVP_air and qRT-PCR was lower and required a higher Ct value threshold of 40 (Spearman r=0.283, p-value=0.031). These data further suggest that the Flu A target in the RVP_air assay required improvement.

### Development of RVP_air_flu: improved detection of Flu A with crRNAs targeted to the Matrix (M) gene

While detection of Flu A in the RVP_air assay corresponded to the same timeframe that Flu A was detected via qRT-PCR (November to March, Fig. 3B), far fewer samples were positive for Flu A in the RVP_air assay compared to qRT-PCR, thus leading to poor sample-to-sample concordance (Fig. 3B, Supplementary Table 1). A BLAST search of the RVP_air Flu A crRNA target indicated that the crRNA was designed to target the A/Thailand/CU-B1512/2016 (H3N2) PB1 gene, and the predominant circulating strain of Flu A for the 2023/2024 respiratory virus season in the region was H1N1 (https://www.cdc.gov/fluview/index.html). In contrast, the qRT-PCR assay targeted a highly conserved region of the Flu A M gene that can amplify both H1N1 and H3N2 targets. Using the ADAPT program, we designed new crRNAs targeting both highly conserved and less conserved regions of the M gene for recently circulating H1N1 and H3N2 viruses. We tested and identified two new crRNAs for H1N1 and H3N2 targets, using A/Wisconsin/67/2022 (H1N1) and A/Darwin/9/2021 (H3N2) M gene targets. These strains were selected based on sequence availability of the M gene and on the inclusion of these virus strains in the 2023/2024 influenza vaccine (23) (https://www.who.int/publications/m/item/recommended-composition-of-Influenza-virus-vaccines-for-use-in-the-2023-2024-northern-hemisphere-Influenza-season).

The new panel focused on influenza, which we named RVP_air_flu. It contained one crRNA that was designed for nucleotides 101-189 of the M gene of A/Wisconsin/67/2022 (H1N1) (Fig. 4A). This region of M has close sequence homology to the same region of the M gene in A/Darwin/9/2021 (H3N2), with only one nucleotide being different between the two viruses (Fig. 4A, left). We also tested a second crRNA designed to nucleotides 453-547 of the M gene of A/Darwin/9/2021 (H3N2). This crRNA has 100% sequence identity to this virus and eight nucleotide differences compared to H1N1 (Fig. 4A, right). We performed the RVP_air_flu assay using *in vitro* transcribed positive control RNA comprising the first 630 nucleotides of the H1N1 and H3N2 gene targets, as well as on RNA isolated from inactivated H1N1 and H3N2 viruses (Fig. 4B). We observed that the crRNA designed to target nucleotides 101-189 of A/Wisconsin/67/2022 (H1N1) could actually amplify both H1N1 and H3N2 positive controls and inactivated viruses (Fig. 4B); thus, we renamed this crRNA pan_FluA. The other crRNA designed to target nucleotides 453-547 of A/Darwin/9/2021 (H3N2) was specific for the H3N2; thus, we called this crRNA H3N2. We re-tested a subset of the samples using these new Flu A crRNAs from the 2023/2024 school year from Fig. 3, emphasizing those from December 2023 to March 2024 (Fig. 3A). Of 143 samples, nine were positive for either pan_FluA or H3N2, compared to 13 samples positive for Flu A via qRT-PCR (Supplementary Table 3). The positive samples detected with RVP_air_flu did not match the samples detected as positive via qRT-PCR very frequently (only 2 concordant samples out of the 13 qRT-PCR positives; Supplementary Table 3). However, the greatest frequency of positive samples for both assays were detected within the same timeframe (November 2023-March 2024; Fig. 4C) and sometimes one assay or the other picked up Flu A genetic material during the same sampling period of 3-4 days, but from different schools (Fig. 4C; Supplementary Table 3). While the concordance was still low, this was still an improvement over the RVP_air Flu A PB1 target, which was only detected in one sample and had no concordance with qRT-PCR positive samples.

**Fig. 4.**
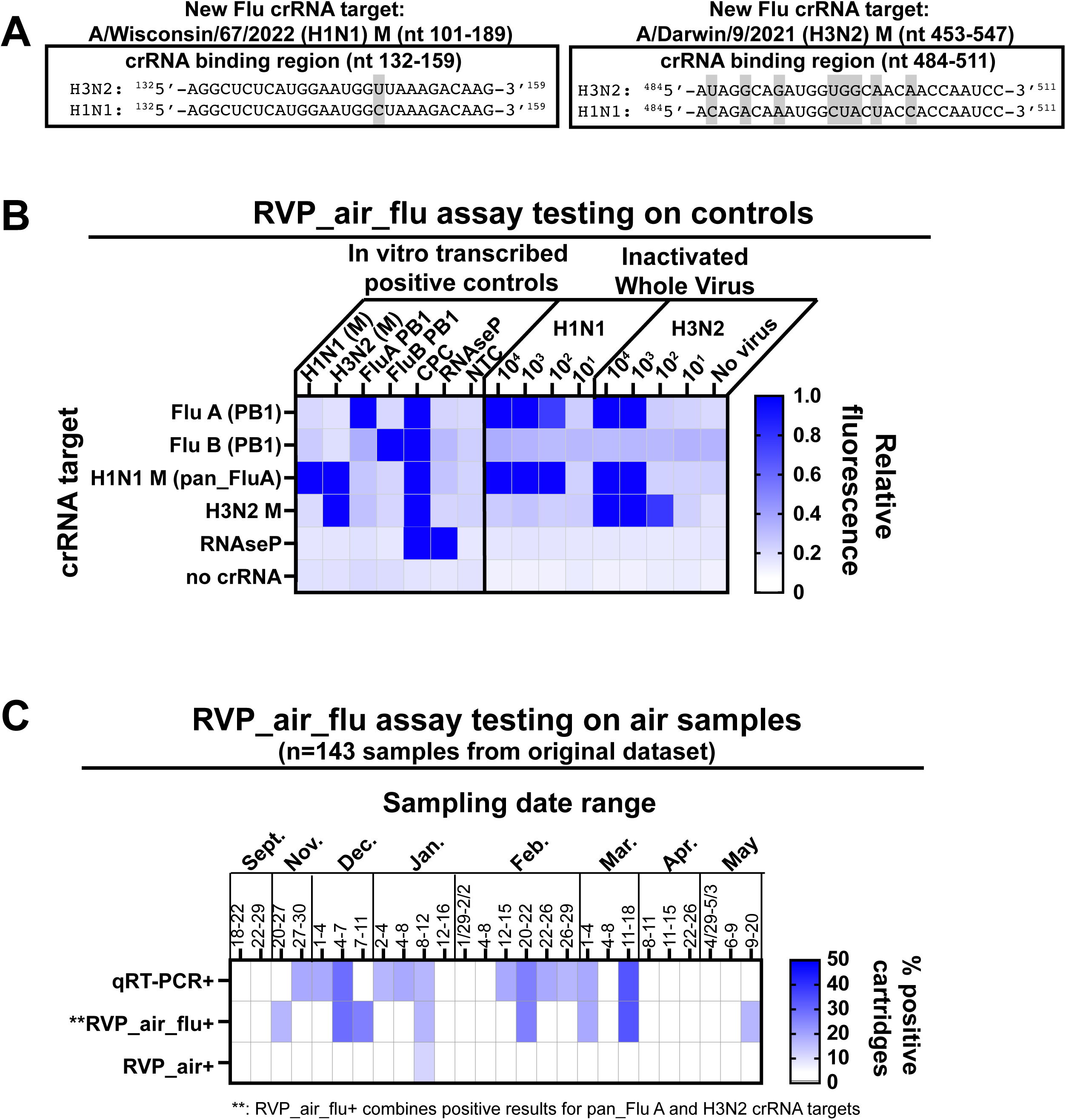
Design of the RVP_air_flu assay. A, To improve detection of influenza A (Flu A), new crRNAs and primers were designed to target nucleotides (nt) 101-189 of A/Wisconsin/67/2022 (H1N1) M gene (left) and nt 453-547 of A/Darwin/9/2021 (H3N2) M gene (right). The boxes on the left and right show the crRNA binding regions for each respective crRNA. Gray boxes indicate nucleotide differences between the H1N1 and H3N2 target sequences within the crRNA binding region. B, RVP_air_flu assay that included the indicated crRNA targets (left) was performed on in vitro transcribed positive controls or on RNA isolated from serial dilutions of inactivated H1N1 (A/California/04/2009) and H3N2 (A/Brisbane/10/2007) stocks. Heatmap colors indicate relative FAM fluorescence intensity of the fluorescent probe used for assay readout. C, The RVP_air_flu assay was performed on RNA isolated from 143 air samples previously run in the RVP_air assay. Comparative heatmaps show the percentage of samples positive for the indicated Flu A targets for qRT-PCR (top), RVP_air_flu targets (middle), or RVP_air targets (bottom).

When directly comparing concordance for both positive and negative results between RVP_air_flu and qRT-PCR for Flu A, there was a moderate improvement in kappa scores compared to the RVP_air panel (RVP_air target alone compared to qRT-PCR, k=-0.029 to 0.085, RVP_air_flu/RVP_air targets compared to qRT-PCR, k=-0.033 to 0.141; Supplementary Table 2). Furthermore, we found a moderately improved correlation comparing the percentage of weekly positive samples for RVP_air_flu targets to qRT-PCR. The most significant correlation was when either RVP_air or RVP_air_flu targets were positive for Flu A and at least one replicate of qRT-PCR for Flu A was below a Ct of 40 (Spearman r=0.399, p=0.026; Fig. 4C).

### Comparison of pathogen genetic material to known infections in the school community

The air samples from the seven schools used for testing by the RVP_air and RVP_air_flu panels are from a single district in Dane County. This district has participated in the Oregon Child Absenteeism Due to Respiratory Disease Study (ORCHARDS) program since 2014 (24). The program was designed to monitor influenza-like illnesses (ILI) in the school district and assess their relationship to absenteeism from school. Nasal swabs were collected from the ORCHARDS participants from September 2023 to May 2024, and testing was performed using the CDC’s influenza and SARS-CoV-2 multiplex assay as well as the Luminex® NxTAG® Respiratory Pathogen Panel. For each indicated sampling period (each time bin was on average 3-4 days), nasal swab samples were considered positive when at least one sample was positive for the targets in the panel (blue color, Fig. 5A).

**Fig. 5.**
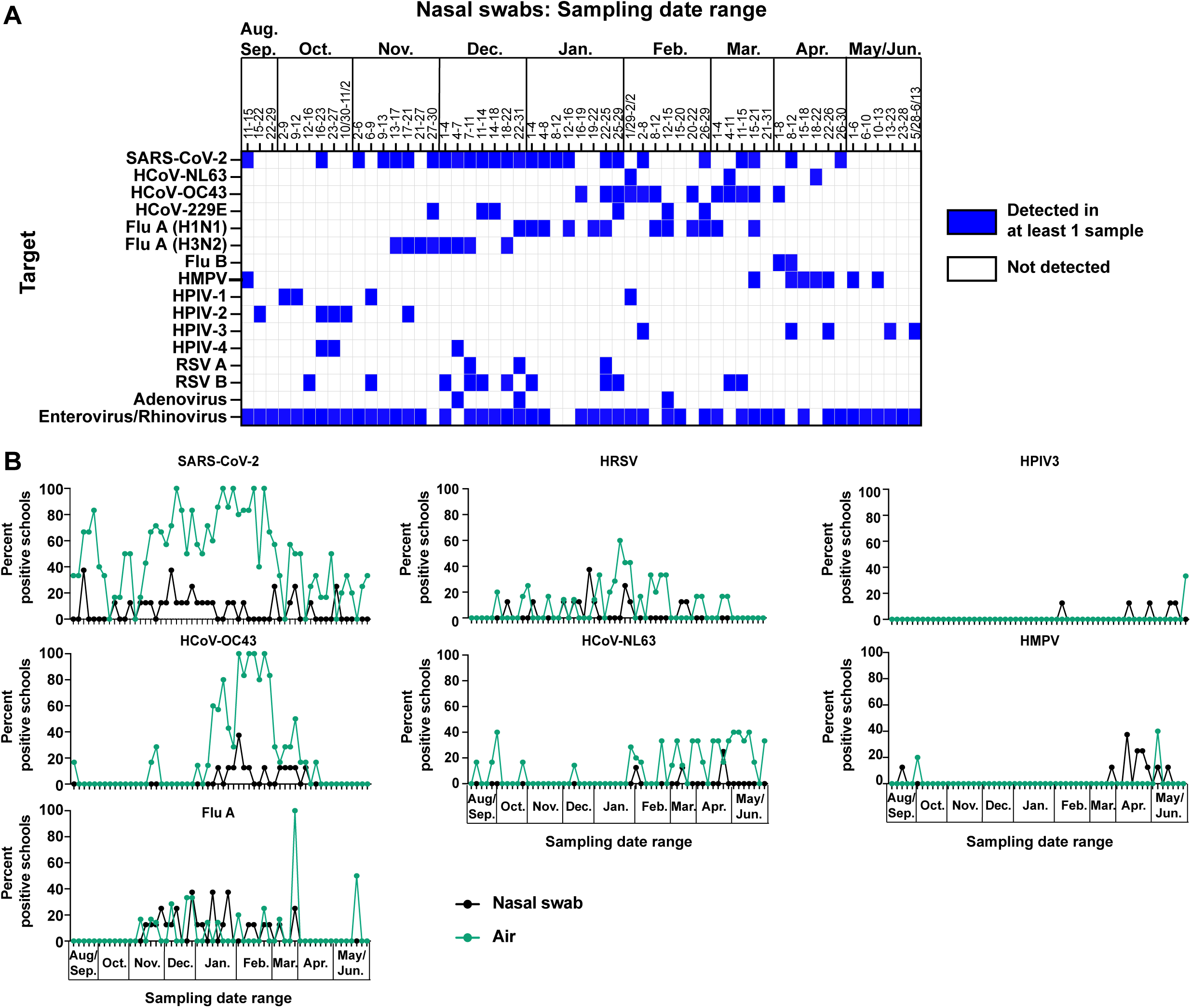
Similar pathogens are detected in air samples and nasal swabs collected from the same K-12 schools. A, Nasal swabs were collected from the same schools where air samplers were placed for the 2023/2024 school year. Nasal swabs were tested in the Luminex® NxTAG® Respiratory Pathogen Panel (RPP) at the Wisconsin State Health Laboratory as described in the methods. The heat map shows sampling periods where at least one nasal swab was positive (blue color) for the pathogens indicated on the left-hand side. B, Nasal swab data shown in Fig. 5A and air sample cartridge data shown in Fig. 3A were recalculated to determine the frequency of schools positive for the indicated pathogens. The percent of schools positive for the indicated pathogens for each sampling time period are shown for nasal swabs (black lines) or air samples (green lines).

We next compared the frequency of schools from which nasal swabs were positive for respiratory pathogens in the ORCHARDS program to the frequency of schools whose air was positive for the same pathogens identified by the RVP_air and RVP_air_flu panels (Fig. 5B). The pathogens targeted for in the Luminex® NxTAG® Respiratory Pathogen Panel assay used on the nasal swabs did not always match the pathogens targeted for in RVP_air and RVP_air_flu assays; Table 7 shows the pathogens used for comparative analysis. Overall, we observed different patterns of relationships between the nasal swab and air. For SARS-CoV-2, Flu A, and HCoV-OC43, pathogens were detected in both air and nasal swabs during similar sampling timeframes. We observed that these pathogens were detected in the air at similar or much greater levels than in the nasal swab data. HRSV and HCoV-NL63 were detected in air samples during time periods when nasal swabs were positive, but also when there were no positive nasal swabs. Finally, for HPIV3 and HMPV, it was difficult to discern a clear relationship because there were limited detections in both the air as well as in the nasal swabs. However, preliminary observations of this limited data showed more frequent detection in nasal swabs compared to air samples.

We performed statistical analysis to determine if there were significant correlations between the increasing and decreasing frequencies of detections between the air and nasal swab data. Significant correlations were observed between nasal swabs and air samples for the ts=0 time bin (i.e., comparisons made within coincident time bins for both nasal swabs and air) for SARS-CoV-2 (Spearman r=0.267, p-value=0.043) and Human Coronavirus OC43 (HCoV-OC43; Spearman r=0.629, p-value<0.001) (Supplementary Fig. 1, ts=0). Lag and lead statistical analysis showed that Flu A and HCoV-NL63 had moderate correlations at negative time shifts (Flu A: ts=-1, Spearman r=0.317, p-value=0.016; HCoV-NL63: ts=-1, Spearman r=0.304, p-value=0.021), while SARS-CoV-2 showed a moderate correlation at positive time shift (SARS-CoV-2: ts=2, Spearman r=0.280, p-value=0.036). HCoV-OC43 exhibited consistently high correlations across all time shifts. When the data were smoothed using a moving average over three time bins, most pathogens reached significant correlation for the ts=0 time bin comparison, with the highest correlation observed for HCoV-OC43 (Spearman r=0.827, p-value<0.001; Supplementary Fig. 1). Overall, we concluded that for pathogens common to both RVP_air/RVP_air_flu and Luminex® NxTAG® Respiratory Pathogen Panel, detection of pathogens in the air by the RVP_air assay could serve as a proxy for the likelihood that there are infected individuals in the corresponding congregate space.

## DISCUSSION

As the use of air surveillance as a low-cost alternative to individual testing in congregate spaces grows, there is a need for similarly low-cost multi-pathogen detection methods to characterize the genetic material of respiratory pathogens in the air. These techniques must be accurate and sensitive within air samples while also generating data that serves as a reliable proxy for circulating infections. In this study, we adapted a multiplexed CRISPR-Cas13 CARMEN RVP assay, which was initially designed for nasal swabs, to air samples collected in K-12 school settings (RVP_air and RVP_air_flu, Figs. 1 and 4). We also took advantage of ongoing respiratory virus testing in one school district to determine whether pathogen genetic material in the air was an indicator of pathogen spread in the community (Fig. 5).

Multi-pathogen detection from environmental samples, such as air samples, is not trivial. The low biomass input and presence of inhibitors hampered pathogen detection when using the original CARMEN RVP assay designed for nasal swabs (Fig. 1; (15)). The iterative approaches we took to redesign crRNAs and remove inhibitors substantially increased the detection of pathogen genetic material from the same samples (Fig. 1). In contrast to nasal swabs where there are a small number of pathogens at a high abundance, air samples may contain a smaller amount of viral genetic material, but that genetic material is likely derived from many individuals shedding distinct viruses. Competitive inhibition between reagents or targets in this mixture will impact assay success (25–27) and could lead to jackpot amplification (28). By modifying the crRNAs and removing inhibitors, we created an environment more suitable for pathogen detection from the air using the CARMEN RVP technology (Fig. 1). Despite these improvements, there were still limitations to detection of pathogens compared to qRT-PCR, where RVP_air and RVP_air_flu could only detect SARS-CoV-2 and Flu A in a smaller proportion of samples with low-to-moderate sample-to-sample concordance. Optimization of every target within the RVP_air and RVP_air_flu panels similar to efforts here for SARS-CoV-2 and Flu A would be required for maximum sensitivity, and even then may not be fully achievable, as was observed for Flu A (Fig. 4). Some of these limitations and observations of lower concordance are also related to the low biomass of air samples. There may be ways to improve the sample processing pipeline, such as the addition of detergents to buffers used for elution which has been successful for increasing the yield of genetic material from environmental samples (29). Overall, with air surveillance, there is a balance between sensitivity of detection, assay throughput, and maximum number of targets for detection that may be difficult to reach with one detection system alone. Our laboratory currently is pursuing other assays, including probe-based multi-pathogen panels and metagenomic sequencing, for pathogen detection to determine the best system that maximizes sample throughput and sensitivity while maintaining relatively low costs.

A key finding from this study was the relationship between the pathogens detected in the air and those detected in children who exhibited influenza-like illness (ILI) and participated in the ORCHARDS program (Fig. 5). We found that the patterns of pathogens detected in the air samples by the RVP_air and RVP_air_flu assays were often concordant with the patterns of detection of the same pathogens from nasal swabs within the school district (Flu A, SARS-CoV-2, HCoV-OC43, Fig. 5B). These findings indicate that air sampling can serve as a reasonable proxy for clinical nasal swab testing in a community. Together, these data further support the argument that effective air sampling and detection of pathogens from K-12 settings can provide a reliable method to detect a pathogen outbreak independent of participation in nasal swab testing, thus giving facilities additional warning to implement mitigation strategies that may reduce disease burden in the community. For example, other studies by the ORCHARDS group found that scheduled school breaks (i.e., breaks of a few days during respiratory viral season) can reduce the spread of ILI (30). Implementing these mitigation strategies at the first sign of increased ILI may limit widespread viral transmission among the individual K-12 community.

There were some differences in the timing and frequency of detection of pathogens in nasal swabs compared to air samples that warrant further investigation. Lag and lead statistical analysis showed that some viruses that were detected in the air prior to detection in nasal swabs (Flu A, HCoV-OC43, HCoV-NL63; Supplementary Fig. 1). This supports the observations by Geenen et al that viral RNA was detected in air samples before detecting the same viruses in facial tissues in a childcare setting (7). A common trend was that pathogens were detected at higher frequencies in the air compared to nasal swabs when both were positive in the same sampling period (SARS-CoV-2, HCoV-OC43; Fig. 5B). For some sampling periods, pathogens were detected in the air but not in nasal swabs (SARS-CoV-2, HRSV, HCoV-NL63; Fig. 5B). One potential reason for these observations is that children may be infected and shedding virus but may not be visibly or symptomatically ill; or alternatively, the families with infected children may be choosing not to participate in clinical nasal swab testing. The ORCHARDS nasal swab data reflects data from individuals that choose to volunteer in the program that are absent from school or visiting nurses’ offices at school due to illness (24). Some viruses, for example SARS-CoV-2 and HRSV, can present frequently with only mild symptoms or subclinical infections in children (31–32), and therefore could potentially still be detected in the air while not being a leading cause of absenteeism/nurse’s office visits. Finally, lag and lead analysis indicated that SARS-CoV-2 increased in the nasal swabs before increasing in the air samples. This trend was mainly evident in the smoothed data (Supplementary Fig. 1). The interpretation is complicated by the fact that SARS-CoV-2 was already present in 20-40% of the air samples prior to when the nasal swab detection began to increase (Supplementary Fig. 1, Fig. 5B). Overall, more sampling data would be needed to determine the exact biological causes of these observations.

CRISPR/Cas-based PCR was developed for rapid, highly sensitive detection of pathogens during the COVID-19 pandemic as an alternative to “gold standard” qRT-PCR (33–34). The CRISPR/Cas system is most noted for being highly sequence-specific, which can minimize off-target effects when used for gene editing (reviewed in (35)). However, one downside to this specificity is that target sequences harboring mutations compared to the crRNA sequence are not recognized for Cas-mediated cleavage. We do not currently know how many mutations can be present while still allowing for Cas-mediated cleavage to occur. The crRNA we designed for H3N2 had eight nucleotide differences compared to H1N1, and could not recognize H1N1 viral genetic material (Fig. 4). In contrast, the crRNA we designed for H1N1 had one nucleotide difference compared to H3N2 and could recognize genetic material from both H1N1 and H3N2 viruses (Fig. 4). Because optimal Cas13 enzymatic activity is highly dependent on the abundance of the target and the crRNA being able to bind with nearly 100% sequence identity to the region of interest, the use of the H3N2 PB1 gene target included in the original CARMEN RVP assay (15) was not optimal for maximum detection of Flu A on air samples (Figs. 3 and 4). For air sampling, where there may be a low abundance of targets and the diversity of target sequences may be great, the design of crRNAs corresponding to the most highly conserved regions within each viral target must be considered for the most efficient use of the RVP_air assay.

While previous studies showed that CARMEN RVP and qRT-PCR had similar sensitivity for detection of pathogens from clinical nasal swabs (15), we observe here that with air samples, qRT-PCR had a greater frequency of SARS-CoV-2 and Flu A detection than RVP_air or RVP_air_flu (Fig. 1, Fig. 3, data not shown). This is likely attributed to several factors, including limited amounts/poor quality of target genetic material present (36–37), different target sequences, differential target amplification efficiency with multiplex PCR compared to single-plex PCR (25–27), and potential differential sensitivity of the Cas13 and Taq polymerase enzymes to residual PCR inhibitors (8, 22, 38). Despite these factors, the RVP_air and RVP_air_flu data were still highly concordant with several pathogens detected in clinical swabs from infected individuals within schools (Fig. 5), indicating that these assays can accurately detect genetic material from respiratory pathogens in the air that are also affecting individuals within a similar timeframe. Overall, the appropriate levels of assay sensitivity and threshold positivity for detecting pathogens in air surveillance studies may vary depending on the surveillance situation, type of pathogen being studied, and quality of the sample.

Compared to wastewater surveillance, air sampling has many unique advantages and can be complementary. Air surveillance can be much more portable and easier to establish than wastewater surveillance. While wastewater sampling can be valuable for community-level surveillance, it does not provide data within individual community settings (schools, emergency departments, long-term care facilities). Also, the predominant pathogens detected in the air are more likely to be respiratory, whereas wastewater sampling is particularly beneficial for detecting GI pathogens. By utilizing air and wastewater surveillance methods, we can better understand the full spectrum of pathogens present at given times within a community. Overall, the two types of surveillance can synergize to provide information beneficial for informing public health.

Even though this study and others (5, 7, 10–11) demonstrate that air surveillance for respiratory pathogens has promise as a cost-effective tool for improving public health, limitations remain. Some limitations are known, such as the efficiency of different air samplers (39) and the limited occupancy within a single location. However, the bigger concern is how to rapidly and accurately detect the maximum number of distinct pathogens in the most cost-effective way. This study demonstrates adapting an inexpensive, high-throughput CRISPR-Cas13 method for pathogen detection of samples containing low biomass and PCR inhibitors. These same challenges apply to the metagenomic sequencing of viruses from air samples to further expand pathogen detection (40–41). The successes in methodological improvement described here provide further confidence that air surveillance collection and detection tools will continue to evolve to improve the molecular assays needed to characterize genetic material collected from air samples.

Ultimately, our study suggests that the modified CARMEN RVP (RVP_air, RVP_air_flu) can be used for routine air monitoring. Further studies of air surveillance in other settings (emergency departments, airports, long-term healthcare facilities) will also help expand our knowledge of how this type of testing may be most beneficial for local communities. A better understanding of the relationship between the detection of pathogen genetic material and transmission risk will further help improve the use of this important technology to improve public health.

## MATERIALS AND METHODS

### Study design and sample collection

ThermoFisher AerosolSense™ air samplers were continuously run for an average of 3-7 days at a time in fifteen schools in Dane County (Wisconsin) during the 2023/2024 school year. After each sampling period, cartridges were removed according to manufacturer’s instructions and transported to the laboratory for processing. Because air is collected from all the people occupying the space in an environment, no individual person is represented. The Institutional Review Board (IRB) of the University of Wisconsin-Madison Health Sciences has determined that air surveillance work in congregate settings is not human subjects research (10).

### RNA extraction and sample cleaning

Individual substrates from air cartridges were eluted in 500µL of phosphate buffered saline (PBS) per substrate and RNA was extracted from 300µL of sample using a Maxwell^Ⓡ^ RSC Viral total nucleic acid purification kit (Promega, Madison, WI) on a Maxwell^Ⓡ^ 48 instrument following manufacturer’s protocols. Each air sample cartridge contains two substrates. From August 2023-February 2024 one substrate was used for RNA isolations, and the second substrate was frozen. The protocol for RNA isolation changed in February 2024 such that both substrates were eluted in 500µL PBS each and isolated on the Maxwell^Ⓡ^ 48 instrument. Following isolation, the RNA from each substrate was combined.

For data described in Figs. 2D and 3-5, PCR inhibitors were removed from 50-100µL of the extracted RNA using the OneStep PCR inhibitor removal kit (Zymo Research, Irvine, CA) according to manufacturer’s instructions.

### Quantitative reverse transcription PCR (qRT-PCR)

Quantitative RT-PCR (qRT-PCR) was performed on RNA isolated from air samples to detect SARS-CoV-2, influenza A (Flu A), and human control RNase P genetic material using a TaqMan Fast virus 1-step kit (Thermo Fisher, Waltham, MA) according to manufacturer’s instructions.

Detection of SARS-CoV-2, Flu A, and RNAseP were performed using protocols previously described in (10, 42). Briefly, previously published primer/probe mastermixes for SARS-CoV-2 nucleocapsid regions and RNAseP (primer/probe sequences from IDT, Coralville, IA) were used for the qRT-PCR and performed on a LightCycler96; cycling conditions are indicated in Table 1.

**Table 1.** SARS-CoV2/RNAseP qRT-PCR cycling conditions.

|  | Temperature | Time | Number of Cycles |
| --- | --- | --- | --- |
| Reverse transcription | 37°C | 2 min | 1 |
|  | 50°C | 15 min | 1 |
| Denature | 95°C | 2 min | 1 |
| PCR | 95°C | 3 sec | 50 |
|  | 55°C | 30 sec |  |
| Cooldown | 37°C | 30 sec | 1 |

Quantitative RT-PCR for influenza A (Flu A) was performed on a LightCycler480 using the following primer/probe sequences (previously tested and described in (43): Flu-A FWD primer; 5’-GGACTGCAGCGTAGACGCTTT-3’, Flu-A REV primer; 5’-CATCCTGTTGTATATGAGKCCCAT-3’, Flu-A probe; 5’-CTMAGYTATTCWRCTGGTGCACTTGCC-3’. Cycling conditions are indicated in Table 2.

**Table 2.** Influenza A qRT-PCR cycling conditions.

|  | Temperature | Time | Number of Cycles |
| --- | --- | --- | --- |
| Reverse transcription | 50°C | 10 min | 1 |
| Denature | 95°C | 20 sec | 1 |
| PCR | 95°C | 15 sec | 50 |
|  | 62°C | 1 min |  |
| Cooldown | 40°C | 30 sec | 1 |

For each air sample, qRT-PCR was performed in duplicate for Flu A and RNAseP targets. For SARS-CoV-2, qRT-PCR was performed in separate wells for each air sample using the primers and probes for the N1 and N2 targets. For all qRT-PCR assays, the average Ct values were determined and viral RNA copies per reaction were calculated based on a standard curve. For each target, a qPCR efficiency within the range of 95%-105% was required for the run to pass; when standards did not pass the run was repeated. For Flu A and RNAseP, the Ct values for duplicates were averaged. For SARS-CoV-2, the Ct values for N1 and N2 were averaged. Positivity was determined as described below in statistical methods.

#### RVP_air and RVP_air_flu assays

A subset of the air samples from seven schools localized to the Oregon school district in Dane county, WI from the 2023/2024 school year was utilized for microfluidic <u>C</u>ombinatorial <u>A</u>rrayed <u>R</u>eactions for <u>M</u>ultiplexed <u>E</u>valuation of <u>N</u>ucleic acids <u>R</u>espiratory <u>V</u>iral <u>P</u>anel (CARMEN RVP) assay analysis and comparisons. The CARMEN RVP assay was performed as previously described (15), with some modifications that are described below, to improve detection of targets with nucleic acids derived from air samples.

#### Design of SARS-CoV-2 nucleocapsid (N) gene and H1N1/H3N2 (M) gene CRISPR RNA (crRNA) and oligonucleotide primer sequences for RVP_air and RVP_air_flu panels

To improve detection of SARS-CoV-2 and Flu A in the CARMEN RVP assay, we designed crRNA and oligonucleotide primers to detect the SARS-CoV-2 nucleocapsid (N) gene (Accession number NC_045512.2) and H1N1 (Accession number OQ203977) and H3N2 (Accession number OQ719010) matrix (M) genes using the Activity-informed Design with All-inclusive Patrolling of Targets (ADAPT) program (Table 3) (20).

**Table 3.**
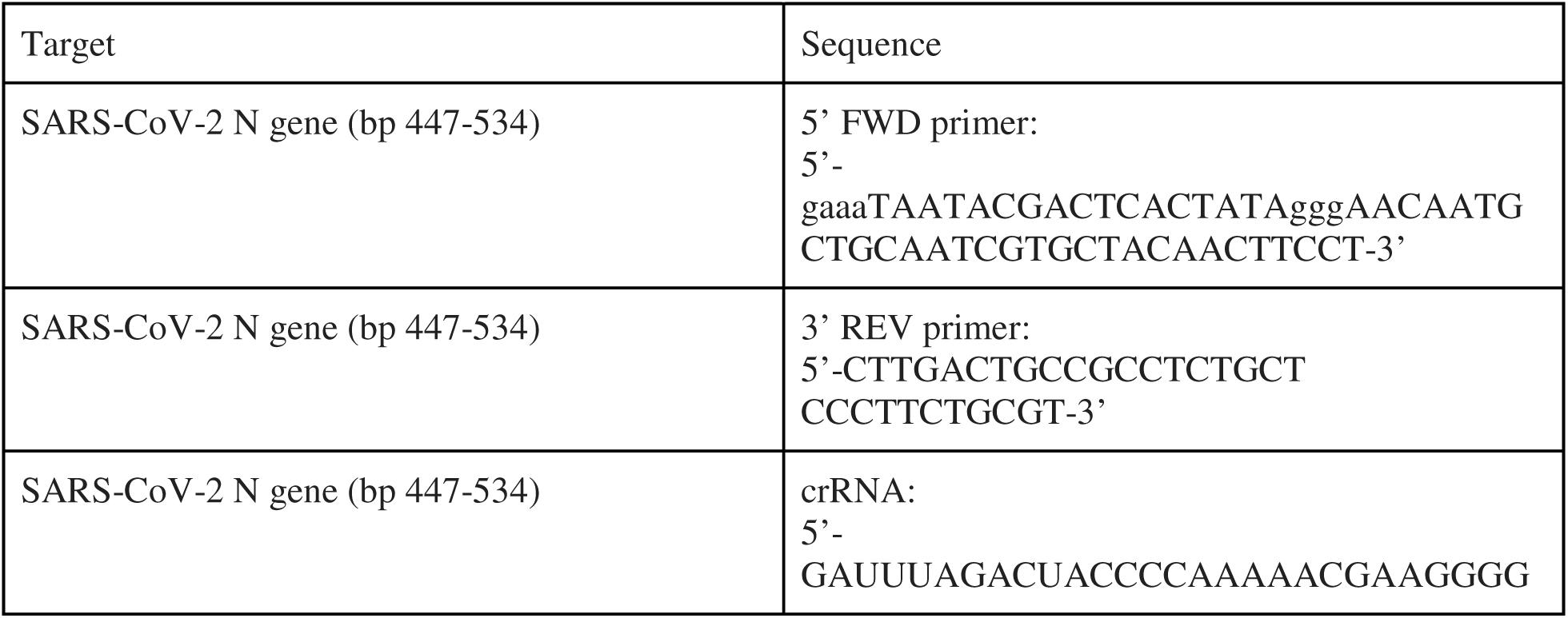
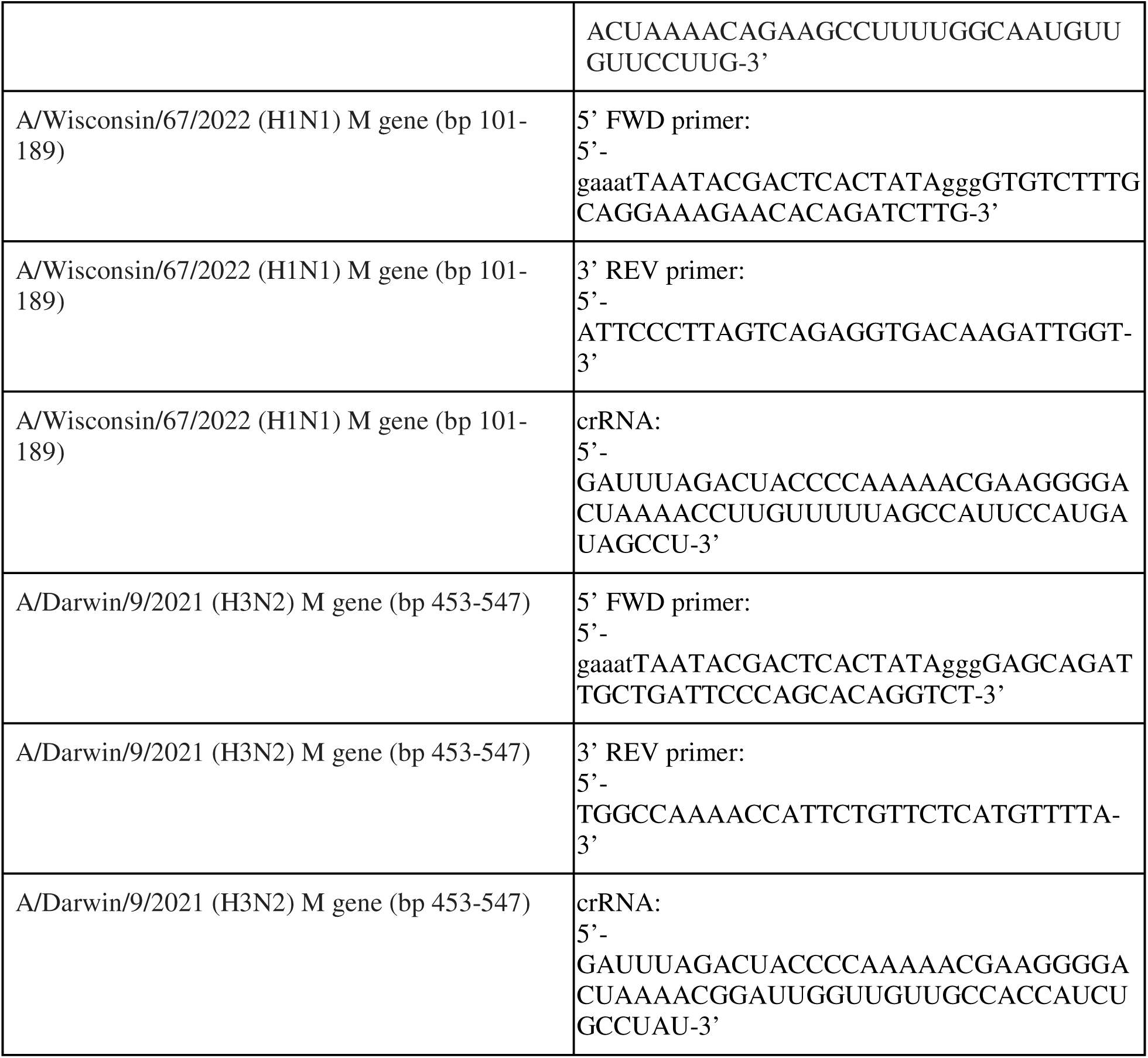
Primer and cRNA to SARS-CoV-2 nucleocapsid (N) and Flu A matrix (M)

#### Reverse transcription and target amplification

Extracted RNA was reverse transcribed and amplified using primer pools to the indicated pathogens (RVP_air; Table 4 and RVP_air_flu, Table 5; Supplementary Table 4) with the Qiagen One Step RT-PCR kit (Qiagen, Germantown, MD) according to manufacturer’s instructions. Other primer/crRNA sequences were previously described in (15), but are also provided in Supplementary Table 4. Samples for both RVP_air and RVP_air_flu were run on a thermocycler using the conditions indicated in Table 6. Cycling conditions were modified from the original CARMEN RVP from 40 cycles to 50 cycles to improve detection of targets from air samples, and primer pool concentrations were also modified to optimize detection of targets from air samples (Table 4; data not shown). RT-PCR was performed in duplicate for each sample.

**Table 4.**
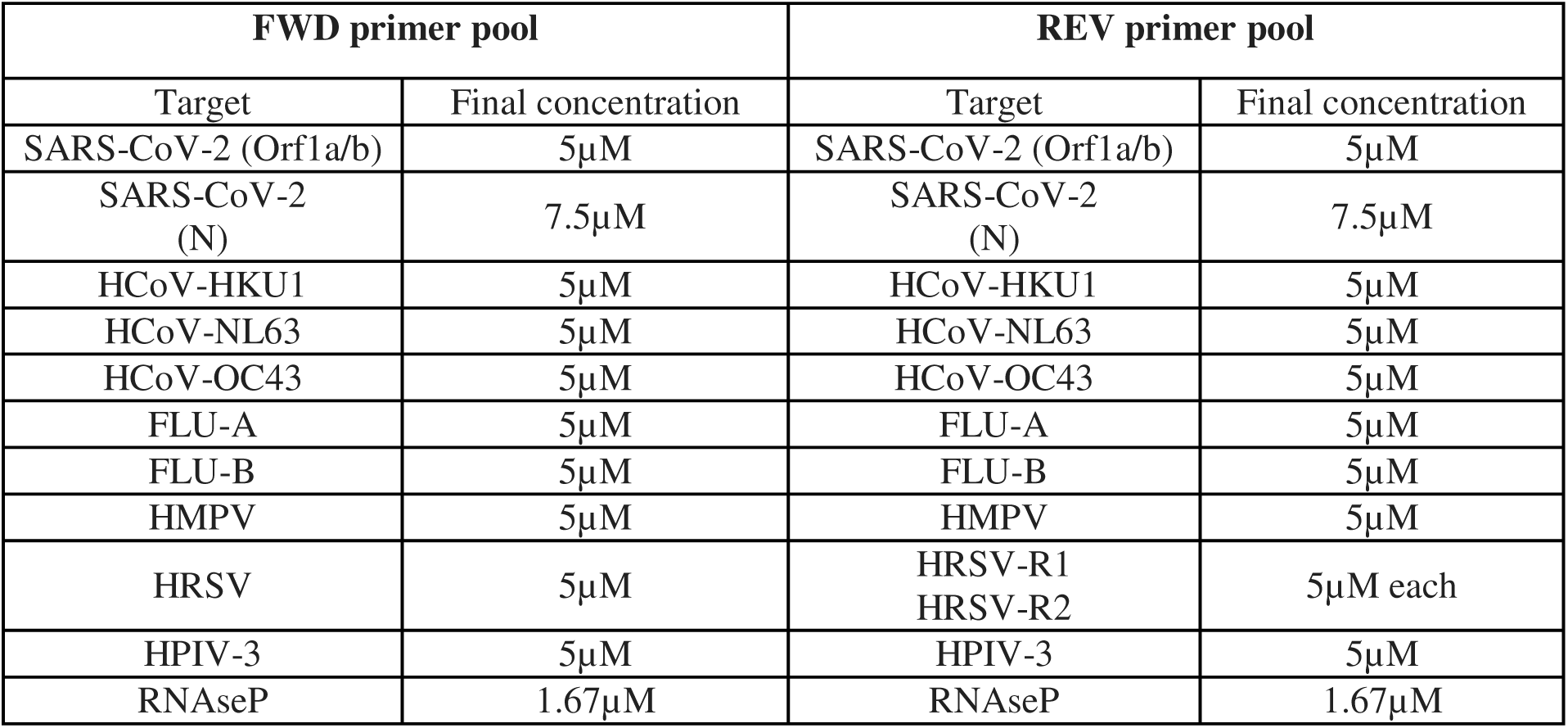
RVP_air primer targets and concentrations.

**Table 5.** RVP_air_flu primer targets and concentrations.

| FWD primer pool |  | REV primer pool |  |
| --- | --- | --- | --- |
| Target | Final concentration | Target | Final concentration |
| FLU-A (PB1)--from RVP_air panel | 5μM | FLU-A (PB1)--from RVP_air panel | 5μM |
| FLU-B (PB1)--from RVP_air panel | 5μM | FLU-B (PB1)--from RVP_air panel | 5μM |
| FLU-A (M; pan_Flu A) | 5μM | FLU-A (M; pan_Flu A) | 5μM |
| FLU-A (M; H3N2) | 5μM | FLU-A (M; H3N2) | 5μM |
| RNaseP | 1.67μM | RNaseP | 1.67μM |

**Table 6.**
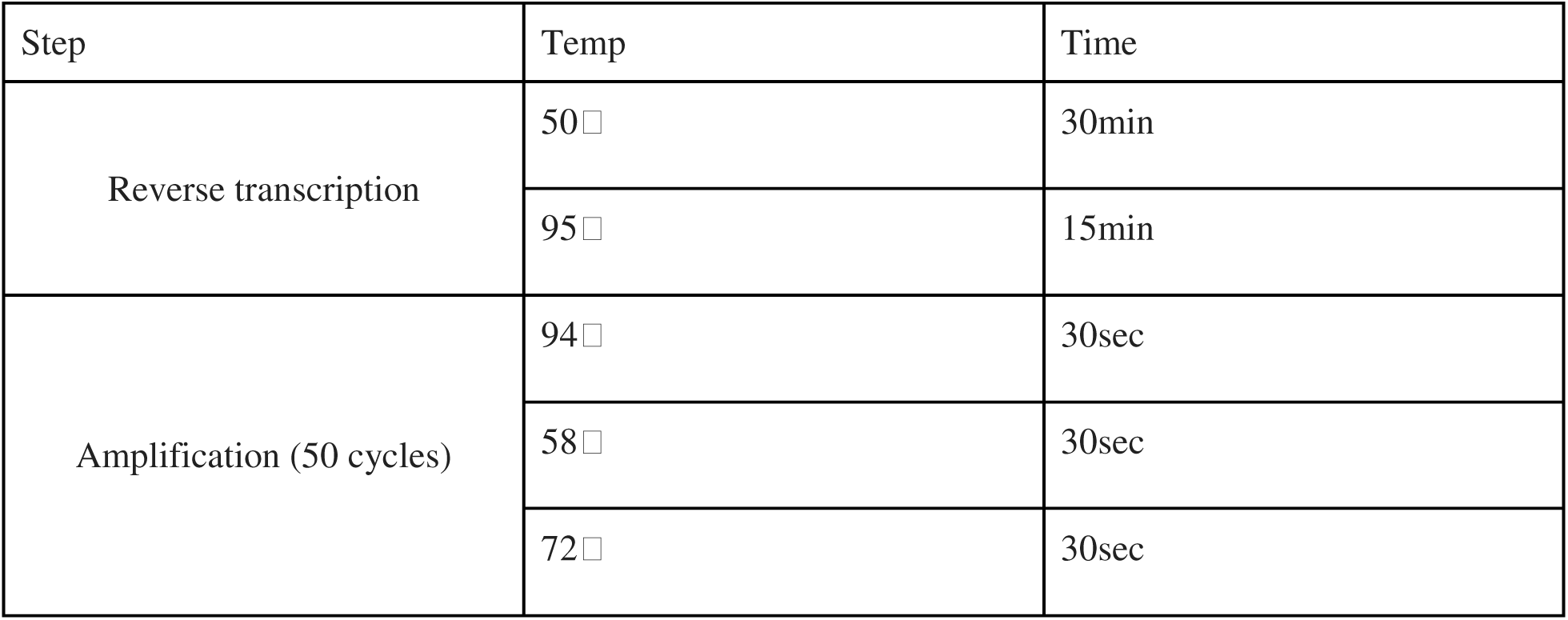
RT-PCR Amplification conditions.

#### Sample and Assay mastermix preparation for the RVP_air and RVP_air_flu assays

RVP_air: Sample and assay mastermixes for loading onto the 192.24 array (Standard Biotools, South San Francisco, CA) were prepared as previously described (15). After preparation of sample and assay mastermixes, samples and assays were loaded onto a 192.24 array according to the manufacturer’s instructions. CrRNA assays corresponding to the 11 RVP_air targets (Table 4) and “no crRNA” control were loaded in duplicate into the 24 assay loading wells of the 192.24 array. RVP_air_flu: Sample and assay mastermix preparation was performed as described above. For loading RVP_air_flu assays onto the 192.24 array, crRNA assays corresponding to the targets amplified by primers described in Table 5 and “no crRNA” controls were loaded in quadruplicate in the 24 assay loading wells.

#### Controls

For each 192.24 array, at least one of each of the following controls was included, similar to controls described in detail in (15): *<u>Combined positive control (CPC)</u>.* G-blocks (IDT, Coralville, IA) corresponding to the target regions for each pathogen for the RVP_air and RVP_air_flu assays were transcribed into RNA using the HiScribe T7 High Yield RNA synthesis kit (NEB, Ipswich, MA). G-block sequences for original CARMEN RVP targets were previously described in (15), but are also shown in Supplementary Table 4. Additional G-block sequences were designed for H1N1 and H3N2 targets, and all G-block sequences are shown in Supplementary Table 4. In vitro transcribed RNA was treated with DNAse I (Thermo Fisher Scientific, Waltham, MA) and precipitated using phenol/chloroform extraction. Total copies of each purified RNA was determined by quantification on a Nanodrop, followed by calculation of the molecular weight of each transcript, conversion to copy equivalents per microliter, and dilution to 10^3^ copies/µL for each positive control. For the SARS-CoV-2 nucleocapsid positive control, SARS-CoV-2 nucleocapsid gene was cloned into PCR2.1 TOPO vector and transcribed using the T7 High Yield RNA synthesis kit and protocol described above. An equal number of copies (10^3^) of each positive control were combined to make the CPC and used as input for RT-PCR. For RVP_air_flu, inactivated H1N1 (A/California/04/2009) and H3N2 (A/Brisbane/10/2007) stocks were purchased from BEI and RNA was isolated to use as controls to test specificity and cross-reactivity of the newly designed crRNAs. *<u>Negative controls:</u>* There were four types of negative controls utilized for the modified RVP_air and RVP_air_flu assays. These controls are identical to those described in (15) with only one exception (Extraction negative control): *<u>No template control (NTC) and Extraction</u> <u>negative control (EC).</u>* For both of the EC and NTC controls, water was loaded during the RT-PCR step to validate that no contaminants were introduced during PCR amplification. In the original CARMEN RVP assay, the EC control was a clinical sample that was positive for RNAseP but negative for all respiratory viruses in the panel (15); however, since qRT-PCR was performed on the RNA from samples used here prior to use in CARMEN RVP, successful RNA isolation was already confirmed. Therefore, we did not include an EC control, but kept the nomenclature as the software to analyze the data requires inclusion of the EC term (15). *<u>Negative detection control (NDC).</u>*Magnesium chloride is required for the enzymatic activity of Cas13 (44, 45). Prior to the addition of MgCl_2_, one aliquot of sample mastermix was placed in one well of a 96-well plate to serve as the NDC. MgCl_2_ was then added to the sample mastermix to a final concentration of 10mM. *<u>No crRNA controls.</u>* Individual crRNA assays were set up in strip-tubes. Each assay setup in strip-tubes included “no crRNA” controls where water was added to prepared assay mastermix solution rather than 1µM crRNA of interest. Each individual assay was then loaded to the 192.24 array in either duplicate or quadruplicate, to fill the 24 assay wells.

### 192.24 array preparation and run

Prepared samples and assays were loaded into a 192.24 array (Standard Biotools Inc, San Francisco, CA) according to manufacturer’s instructions. The loaded array was run on a Biomark X using a previously optimized custom set of cycling conditions (described in (15)). Data were analyzed using the CARMEN-RVP software program (available and described in (15)) to initially identify positive hits. Samples were run in duplicate, and the relative fluorescence intensity was measured and averaged. Positive hits were validated manually as samples with average relative fluorescence intensity 1.8 times higher than the fluorescence of non-template controls (NTC) present in the same run.

### ORCHARDS sample collection and processing

#### Human subjects statement

All components of the <u>OR</u>egon <u>CH</u>ild <u>A</u>bsenteeism due to <u>R</u>espiratory <u>D</u>isease <u>S</u>tudy (ORCHARDS) were reviewed and approved by the University of Wisconsin Education and Social/Behavioral Science and Health Sciences Institutional Review Boards (protocol 2013-1357). The study is in full compliance with the Health Insurance Portability and Accountability Act of 1996 (HIPAA), Family Education Rights and Privacy Act (FERPA), and all other federally mandated human subjects regulations. The US Office of Management and Budget approved forms used in ORCHARDS.

#### Sample collection and processing

Nasal swab specimens were collected each week as part of the ORCHARDS program (24). All specimens were tested for influenza A/B and SARS-CoV-2 by reverse transcription polymerase chain reaction (RT-PCR) using the CDC’s influenza and SARS-CoV-2 multiplex assay at the Wisconsin State Laboratory of Hygiene (WSLH) as described in (1). All specimens were also tested for multiple respiratory pathogens at the WSLH using the Luminex^®^ NxTAG^®^ Respiratory Pathogen Panel (RPP) as previously described (1, 46). Respiratory virus targets in this RPP included influenza (Flu) A matrix, FluA H1N1, FluA H3N2, FluB, RSV-A, RSV-B, Human Coronavirus (HCoV)-229E, HCoV-OC43, HCoV-NL63, HCoV-HKU1, Human Metapneumovirus, Rhinovirus/Enterovirus, Adenovirus, Human Parainfluenza Virus (HPIV) 1, HPIV-2, HPIV-3, and HPIV-4.

### Statistics

#### RVP_air panel optimization (Fig. 2)

In optimization experiments performed in Fig. 1, to assess agreement among qRT-PCR and the original CARMEN RVP panel (15) and with the modified CARMEN (RVP_air) panel, we performed Cohen’s kappa tests (GraphPad Prism software). These comparisons were limited to comparisons for detection of SARS-CoV-2 and influenza A. The kappa scores (κ) are reported.

#### RVP_air and RVP_air_flu comparisons with qRT-PCR data (2023/2024 samples, Figs. 3-4)

*Determining positivity:* For CARMEN RVP, RVP_air, and RVP_air_flu, assay positivity was determined as described in the methods and in (15). For qRT-PCR Flu A and RNAseP, the Ct value for duplicates were averaged. For SARS-CoV-2, the Ct values for N1 and N2 were determined and then averaged. For the qRT-PCR assay, an average Ct value below 40 was considered positive. For statistical analysis, we applied two different Ct cutoffs to explore which Ct cutoff was best for comparative analyses between RVP_air/RVP_air_flu and qRT-PCR data. A result for either SARS-CoV-2 or Flu A was considered positive if both replicates had Ct values either equal to or below 35 or 40. In the secondary analysis, a result was classified as positive if at least one of the replicates had a Ct value equal to or below 35 or 40.

#### Criterion for primary and secondary analyses

Comparative statistical analyses were performed for SARS-CoV-2 between RVP_air and qRT-PCR (n=347 samples; Fig. 3). For SARS-CoV-2, the primary comparative analysis (qRT-PCR vs. RVP_air; n=347 samples) focused on gene N, but a secondary analysis was also performed in which a result was considered positive if SARS-CoV-2 N or Orf1ab targets were positive. For Flu A, comparative statistical analyses were performed between RVP_air targets and qRT-PCR (n=347 samples) and RVP_air_flu and qRT-PCR (n=143 samples; Fig. 4). For Flu A (qRT-PCR vs. RVP_air/RVP_air_flu), the primary comparative analysis was conducted on the subset of samples that were run using the RVP_air_flu assay (n=143) and a result was considered positive if the Flu A targets in either RVP_air_flu (pan_Flu A and H3N2) or RVP_air (Flu A PB1) were positive. The secondary analysis for Flu A was based on the full RVP_air dataset (n=347).

#### Kappa score calculation

For both the primary and secondary analyses described above, the P-value for the kappa statistic was calculated using a permutation test. In this test, positive and negative result labels were randomly permuted 100,000 times. The P-value was determined by dividing the number of random permutation kappa scores that were equal to or higher than the observed kappa by the total number of permutations.

#### Spearman correlation tests

Spearman correlation tests were performed to evaluate if the percent of positive cartridges by qRT-PCR and RVP_air and RVP_air_flu followed the same trends for each time bin (Fig. 3B-C; Fig. 4C). Spearman correlation tests were also performed to determine if the weekly change in the percent of positive cartridges followed the same trends across multiple time bins (i.e, the difference between two adjacent time bins).

#### RVP_air and RVP_air_flu comparisons with ORCHARDS data (Fig. 5)

ORCHARDS and RVP_air/RVP_air_flu did not include the exact same pathogen panel; therefore, analysis was limited to pathogens similar between both datasets. For each shared pathogen, we conducted a comparative analysis. Specifically, we compared RVP_air/RVP_air_flu and ORCHARDS target results for the pathogens indicated in Table 7:

**Table 7.** Pathogens used for comparative statistical analysis between CARMEN RVP and ORCHARDS data.

| **Pathogen** | **RVP_air and RVP_air_flu targets** | **ORCHARDS targets** |
| --- | --- | --- |
| influenza A | pan_FluA, H3N2, Flu A (PB1) | influenza (Flu) A matrix, FluA H1N1, FluA H3N2 |
| influenza B | Flu B (PB1) | FluB |
| Human coronavirus NL63 | HCoV-NL63 | HCoV-NL63 |
| Human coronavirus OC43 | HCoV-OC43 | HCoV-OC43 |
| Human metapneumovirus | HMPV | HMPV |
| Respiratory Syncytial Virus | HRSV | RSV-A, RSV-B |
| Human Parainfluenza Virus-3 | HPIV3 | HPIV3 |

To align the data between datasets according to sampling time, we binned results from ORCHARDS to match the time bin structure of the air samples used for RVP_air (Fig. 5). For each pathogen separately, if no positive result was observed in a given time bin for any ORCHARDS samples within that bin, the result was classified as negative. A result was considered positive for a given pathogen if one or more ORCHARDS samples were positive within that time bin. To evaluate whether the detection of a pathogen in the air by RVP_air correlates with its detection in participants tested in ORCHARDS, we performed a Spearman correlation between two vectors representing the percentage of positive schools per time bin. If multiple RVP_air results were available for the same school within a given time bin, the pathogen was considered positive if any of the results were positive. As a secondary analysis, we conducted a Spearman correlation using smoothed data by applying a moving average. Specifically, at each time index i, the value was calculated as the mean percentage of positive schools at time indices i, i−1, and i+1. For edge handling, the average was computed over the available two values. Additionally, we performed a lag or lead analysis to assess whether RVP_air detection tended to precede or follow ORCHARDS detection. We evaluated time shifts ranging from -3 to +3 time bins. This analysis was conducted on both the original and smoothed data, as described above.

#### Statistical testing parameters

All tests were conducted at significance level P ≤ 0.05. Permutation test of kappa score was done with one-sided null hypothesis, all other tests were done with two-sided null hypothesis. All analyses were performed in Python 3.12.7 with the package scipy 1.15.1 and sklearn 1.6.1. While we present primarily unadjusted P values in accordance with our prespecified plan, and consistent with our usual practice, we value and encourage consideration of the impact of multiple testing. For post hoc multiplicity adjustment over any set of tests, we provide all unadjusted P values in Supplementary Table 2.

## Data Availability

All data produced in the present study are available upon reasonable request to the authors.

https://drive.google.com/drive/folders/1THUy4ZCXNmK_nR5eEFPgKt6dNMNMdfZc

## ACKNOWLEDGEMENTS

This study was funded through NIH R01 grant 5R01AI170737. We thank the staff and schools within the Dane County, WI school district for participation in the air surveillance program. We also thank the participants of the ORCHARDS program. ORCHARDS was supported by the Centers for Disease Control and Prevention (CDC) through cooperative agreement #5U01CK000542-02-00 and #6U01CK000630-01-01.

